# Less Haste, More Speed: Does delayed blood culture loading lead to adverse incubation time or yield?

**DOI:** 10.1101/2024.10.15.24315399

**Authors:** Gavin Deas, Fergus Hamilton, Philip Williams

## Abstract

**Background:** Blood culture remains a vital diagnostic tool in the acutely unwell patient. The UK Standards for Microbiology Investigations (SMI) stipulate pre-analytical requirements that are aimed at increasing yield and reducing turnaround time. The requirement to load blood cultures onto incubators within 4 hours has shown to reduce turnaround time but limited evidence exists as to whether it improves yield.

**Methods:** We extracted blood culture results including organism growth, time to detection (TTD), location and time of sample collection from 4 hospitals in Southwest England. We then used mixed effects, Bayesian linear and logistic regression models to examine the effect of predictor variables like time to loading (TTL) on the response variable of growth or TTD. We used generalised additive models to explore non-linearity.

**Results:** 449,191 culture sets were analysed, 398,077 of which had enough data to include in the final analysis. 37,255 sets flagged positive (9.36%) of which 21,330 were considered pathogens. Our primary analysis identified a small decrease in yield with each hour delay in loading, OR 0.997 (95% credible interval, CrI 0.994 - 1.001). This effect was largest in *Streptococcus pneumoniae, agalactiae* and *pyogenes*. In our analysis on TTD, culture sets spend 10.98 (95% CrI 7.8 – 13.2) minutes less incubating for each hour delay. There was no convincing non-linearity in either of these effects.

**Conclusion:** There is a marginal loss of growth for every hour a blood culture is left unincubated, with the loss of recovery of *Streptococcus pyogenes* and other streptococci most common. There was no evidence of a reduction in Gram-negatives, anaerobes, or yeasts. There was a small decrease in TTD for delayed sets. This analysis suggests there is only marginal benefit in reducing time to load blood cultures on incubators.

**Highlights:**

- The UK SMI stipulates blood cultures be loaded within 4 hours of being taken
- The effect on yield has not been determined for delayed loading
- We found overall that there was a marginal 0.3% loss of yield per hour delay
- There was a significant loss of *Streptococcus pyogenes* at 3.2% per hour delay

## Background

Blood culture remains a standard and common method for diagnosing bacteraemic patients and is often used in the setting of the acutely unwell, undifferentiated patient groups [1]. Although culture remains the gold standard, it is recognised that it is insensitive, with many patients remaining culture-negative despite clear clinical evidence of infection [2] [3] [4]. Additionally, due to the requirement for culture and identification results are often not available to clinicians for more than 24 hours from sampling. As such, many organisations have set standards for blood culture in an attempt to increase both the speed of turn around (time to positivity, TTP) and yield of blood culture [5].

In the UK, the Standards for Microbiology Investigation (SMI) for Sepsis and Systemic or Disseminated Infections includes recommendations to load samples onto an incubator within 4 hours of collection [6]: the time to load (TTL). This pre-analytical requirement for TTL goes alongside other requirements on the inoculum size with volume requirements and bottle numbers to increase the clinical utility of the test [6]. The evidence supporting the recommendation on TTL includes an effect on total time between collection and pathogen detection (TTP) [7].

Remarkably the effect of reduced TTL on yield is still not clearly understood. It seems plausible that earlier incubation of blood cultures could improve yield, particularly of pathogens that are temperature sensitive. However, some evidence supports pre-incubation as *decreasing* yield, a paradoxical result that has not been widely explored [8]. Although studies [9] [10] have been performed of in-vitro analysis of spiked blood cultures, it is not clear that these analyses reflect the reality of clinical microbiological sampling where microbiological load is often unknown, and patients may have received antimicrobials or other treatments that might alter microbial growth [11]. As such, we performed a large (>300,000 blood culture bottles) analysis on the effect of TTL on a) yield, and b) time to detection.

## Methods

### Setting and study inclusion

This is a retrospective observational analysis of the blood culture pathway in 4 hospitals in the Southwest of England, including 2 district general hospitals and 2 tertiary referral centres. They saw a combined 31,398 patients in March 2024 in their emergency departments and have an overnight bed base of 2,664 [12]. Each site has blood culture (BACTEC FX) incubation machines. All blood culture bottles sent to our laboratory from the 1^st^ January 2017 to 30^th^ June 2024 were analysed. Culture sets are taken in line with local antisepsis protocol at clinician’s decision. Blood culture sets were processed in line with UK SMI 12; briefly bottles are loaded continuously onto automated incubators as soon as they arrived in the laboratory. Fluorescence based automated alerts indicate a positive result and identification of organisms is provided through gram stain, antigenic grouping, matrix-assisted laser desorption ionization – time of flight (MALDI-TOF), and VITEK2 (bioMérieux) automated antimicrobial susceptibility testing.

### Clinical data

We collected information on samples received on a) ward and hospital, b) timing of the blood culture (collection, loading, result, MALDI-TOF result), and c) date. The ICE (Integrated Clinical Environment^TM^) requesting system was used to establish sample collection time. The BD BACTEC FX incubator collects loading time (TTL) and time to detection (TTD) automatically. Duplicate sets of blood cultures were removed, and the fastest TTD was treated as the TTD. Organisms were classified into pathogenic organisms or contaminants by the authors, based on whether they pose clinical significance to patients on a population basis. Although any organism can be considered pathogenic or a contaminant given the right circumstance, we chose organisms that pose significant clinical disease, form the mainstay of clinical microbiological work, and those that the Standards of Microbiological Investigation aim to target [6]. This list is available in supplementary material 1.

### Statistical methods

Descriptive statistics of the sources, TTD and TTL, and organism are provided. Naïve linear and logistic regression analysis, unadjusted and adjusted Bayesian modelling was performed and generalized additive models are used to fit splines to explore the presence of non-linearity. One potential problem with analysing time to loading is that estimates of growth are biased by the physical location of wards and hospitals and whether samples were taking during laboratory operating hours. A ward that we might expect to have a high positivity rate, for example an intensive care unit, being far from the laboratory would produce a paradoxical effect of an increased time to loading correlating with positive culture. To overcome this, we built a mixed effects model that accounted for ward and site, with wards nested within hospitals. Formally, we allowed a random intercept for each ward, nested within each hospital, essentially allowing each ward to have its own baseline positive rate. We then ran logistic regression (for yield) and linear regression (for TTD), assuming a fixed effect for both (i.e. that the effect on yield of delay was the same across wards and sites). There are 8 sites: each of the 4 hospitals were divided into emergency departments and inpatient groups, and there were 107 wards in total. Due to the large number of wards and to allow model fitting, we used a Bayesian model with weakly informative priors. Statistical analysis was performed using R 4.2.2 utilising the CRAN package brms [13]. Akaike Information Criterion values were used to choose optimal model fitting.

The logistic/linear regressions models chosen took the form *(growth ~ TTL + working day + year + (1 | site/ward)* and (*TTD ~ TTL + working day+ year + organism + (1 | site/ward)*, where *working day* was a binary variable of the sample being taken between 0900-1700. This was used on the dataset with all isolates and then subsequently on a subset with pathogenic isolates only. Pathogen specific analysis for the most common isolates (n > 250) and for anaerobic and fungal growth was also performed. The code is supplied in Supplementary Material 2.

### Ethics

All data used in this study was anonymised at source on extraction from the clinical systems. No patient identifiable information was ever extracted, simply the ward, location, time of sample, and whether it was positive (including the organism isolated). As such, ethical approval was not required [14].

## Results

### Descriptive statistics

449,191 sets of blood cultures were collected. This averaged out at 189.45 sets per day and 71.12 sets per 1000 bed days. Entries with data loss for the loading time, time on incubator, organism identification, or polymicrobial culture samples were removed leaving 398,077 sets. 37,255 sets flagged positive (9.36%) of which 21,330 were considered pathogenic and 15,925 (4.0%) contaminants: with 14,752 of the contaminants being coagulase negative staphylococci. The mean time to loading (TTL) was 4.07 Hours (median 2.92 Hrs; Q1 1.35 Hrs; Q3 5.52 Hrs). A density plot of total cultures received is show in Figure 2 broken down by site which clearly shows that the TTL was different across differing sites. This data is also shown in Table 1, alongside data on the average positivity rate at each site.

**Figure 1.**
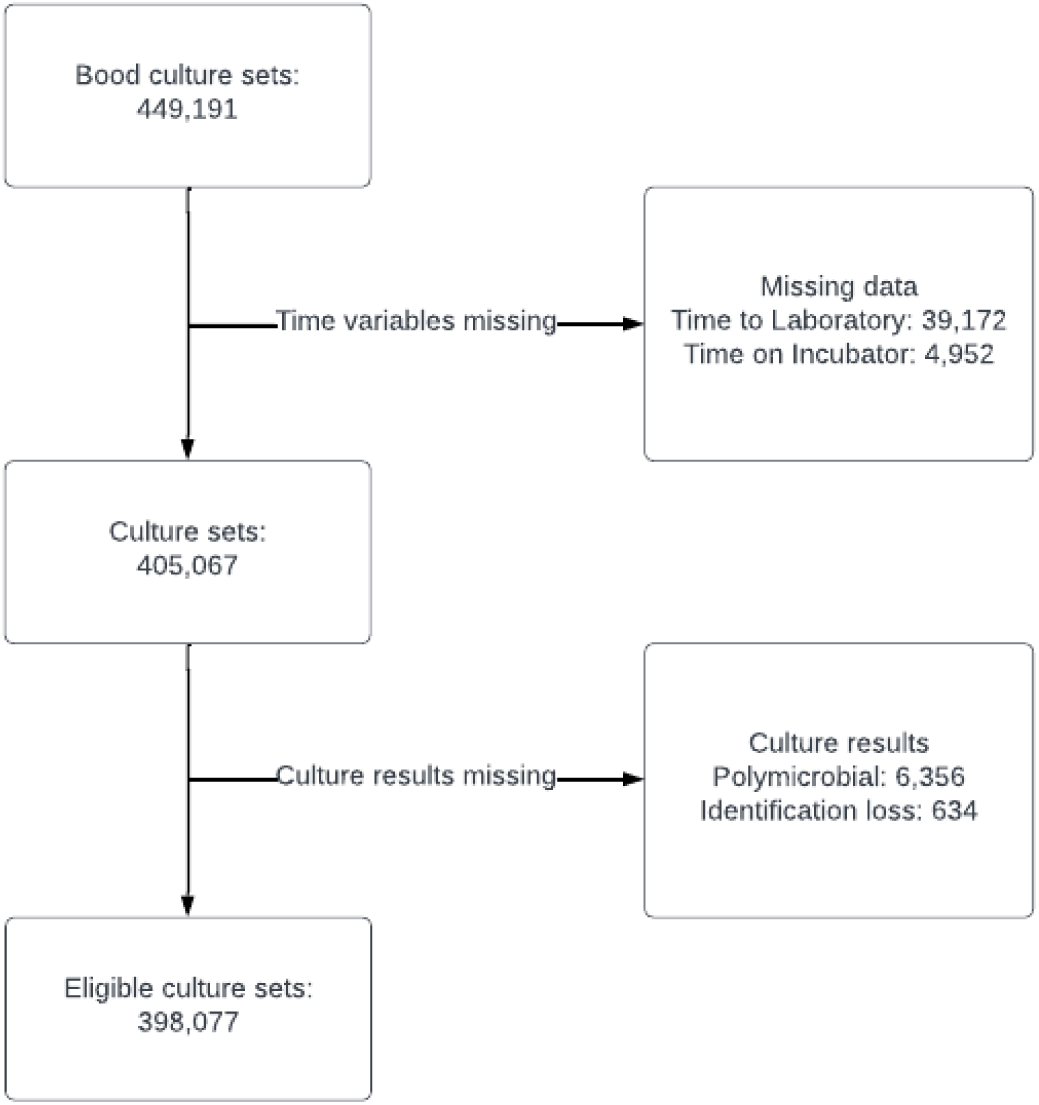
Flow chart of culture set inclusion.

**Figure 2.**
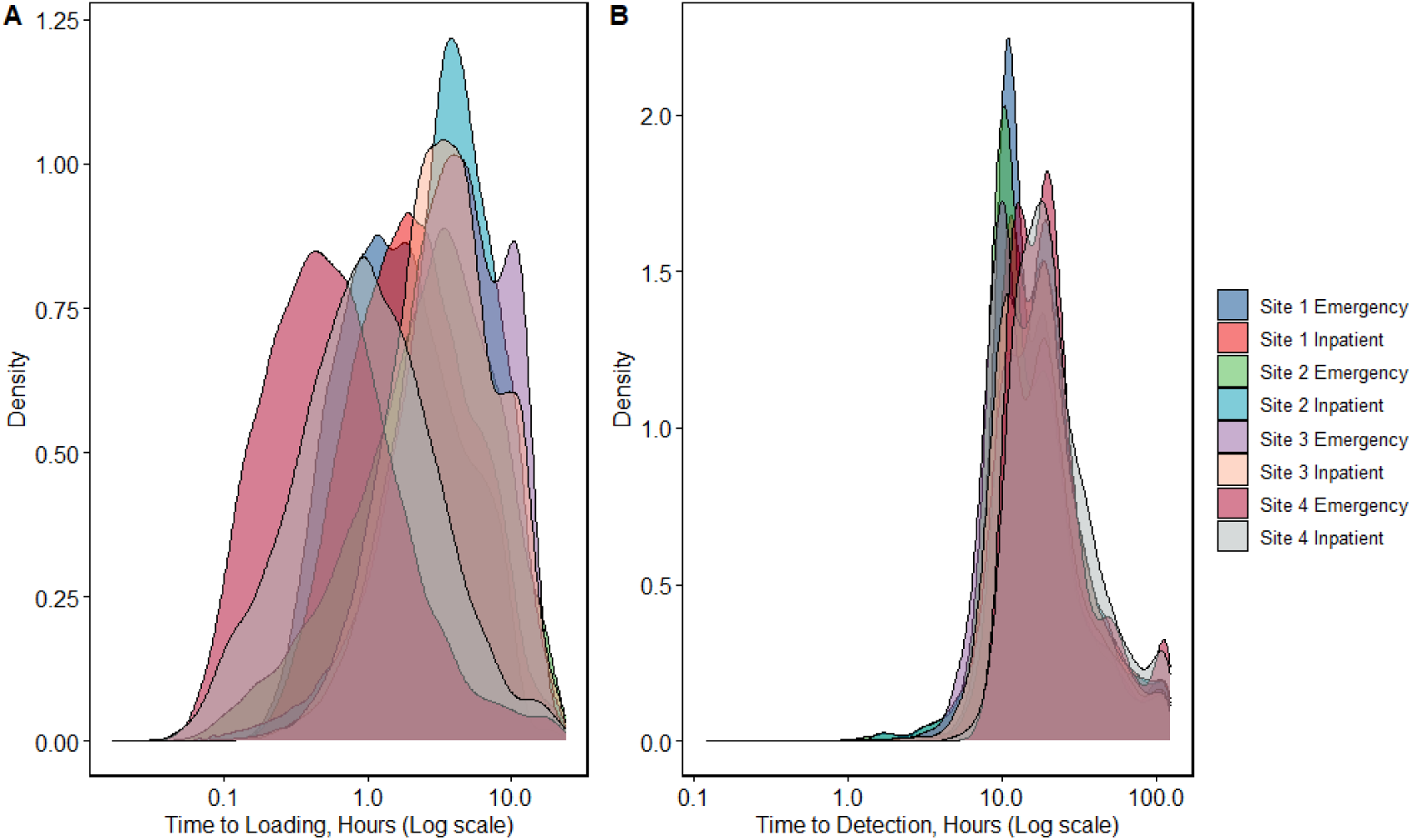
A) density plot showing logarithmic distribution of Time to loading per site. B) A density plot showing time to detection per site.

**Table 1.** Top 25 sources for blood cultures, showing mean, median, standard deviation, 1^st^ and 3^rd^ quantile for the time to loading, number of cultures received and the positivity rate for that site; ordered by TTL.

| Source | Mean TTL | Median TTL | SD | N | Rate |
| --- | --- | --- | --- | --- | --- |
| Site 4 Emergency | 1.15 | 0.50 | 2.25 | 18185 | 0.13 |
| Site 1 Emergency | 2.51 | 1.57 | 2.57 | 22784 | 0.15 |
| Site 1 AAU | 2.98 | 1.95 | 2.90 | 8752 | 0.08 |
| Site 1 AMU | 3.06 | 1.95 | 3.12 | 4881 | 0.08 |
| Site 1 Emergency MIU | 3.62 | 2.60 | 3.25 | 8871 | 0.09 |
| Site 1 Delivery suit | 3.83 | 2.77 | 3.23 | 4090 | 0.04 |
| Site 2 Paediatric Emergency | 3.87 | 2.87 | 3.40 | 13640 | 0.08 |
| Site 3 SAU | 4.06 | 2.82 | 3.61 | 3546 | 0.08 |
| Site 2 Emergency | 4.12 | 2.85 | 4.04 | 22152 | 0.13 |
| Site 2 Cardiac ICU | 4.24 | 3.38 | 3.44 | 5663 | 0.07 |
| Site 1 Nephrology | 4.32 | 2.85 | 4.09 | 3894 | 0.11 |
| Site 2 Medical ward | 4.43 | 3.57 | 3.51 | 3668 | 0.08 |
| Site 3 Haematology | 4.45 | 3.28 | 3.57 | 7063 | 0.1 |
| Site 2 Paediatric oncology | 4.59 | 3.70 | 3.60 | 5609 | 0.07 |
| Site 3 Paediatric | 4.77 | 3.45 | 3.83 | 3411 | 0.07 |
| Site 2 Delivery suite | 4.90 | 4.08 | 3.44 | 5555 | 0.03 |
| Site 2 Neonatal ICU | 4.98 | 4.14 | 3.45 | 3732 | 0.08 |
| Site 3 Emergency | 5.10 | 3.88 | 3.89 | 32008 | 0.13 |
| Site 3 MAU | 5.11 | 3.80 | 4.05 | 5727 | 0.1 |
| Site 2 Haem/oncology | 5.32 | 4.18 | 3.81 | 7458 | 0.07 |
| Site 1 ITU | 5.59 | 4.63 | 3.99 | 13890 | 0.11 |
| Site 2 Haematology | 5.66 | 4.45 | 3.90 | 14211 | 0.07 |
| Site 3 ITU | 6.05 | 4.83 | 4.25 | 6157 | 0.09 |
| Site 3 Emergency MIU | 6.55 | 5.35 | 4.41 | 4400 | 0.13 |

Table 1 highlights the problem with naïve regression without accounting for ward structure. Emergency Departments (ED) have a very high level of positivity, as expected, but in one setting (Site 4 Emergency), the mean TTL is extremely low, as the laboratory is very closely co-located to this ED. A naïve regression analysis of this site would identify this as suggesting a decreased TTL leads to increased yield, but this may just reflect the colocation of this laboratory and ED. In contrast, the ED in Site 3 has the highest TTL and a very high rate of positivity: a naïve analysis here would identify the opposite association here.

### Decreased TTL leads to a marginal increase in yield in both unadjusted regression and in mixed effects models

In an unadjusted regression model, there was strong evidence of a small effect (OR per hour, 0.986 95% credible interval, CrI 0.983 – 0.990). This can be interpreted as a 1.4% decrease in yield for each hour delay. In contrast, our mixed effects model accounting for site level variation had weaker evidence of an even smaller effect, an OR for growth of 0.997 (95% CrI 0.994 - 1.001) per hour: e.g. a decrease of around 0.3% per hour.

We then performed non-linear modelling to show this and identify if there was a threshold effect where yield suddenly decreased. The unadjusted model is shown in Figure 3, and mixed-effect adjusted model in Figure 4. In the unadjusted model, there was clear evidence of non-linearity and even non-monotonicity with a large decrease in yield over the period 0-4hrs, but then almost no effect until ~12hrs, with a subsequent decrease in yield when the TTL increased after 12h. In contrast, when accounting ward level effects and time in the mixed-effects model, we see no evidence of non-linearity nor a threshold effect, and the data was consistent with a small, linear effect in reducing yield. The Akaike Information Criterion (AIC) values were smaller for the model that accounted for covariates in both the linear and non-linear models,

**Figure 3.**
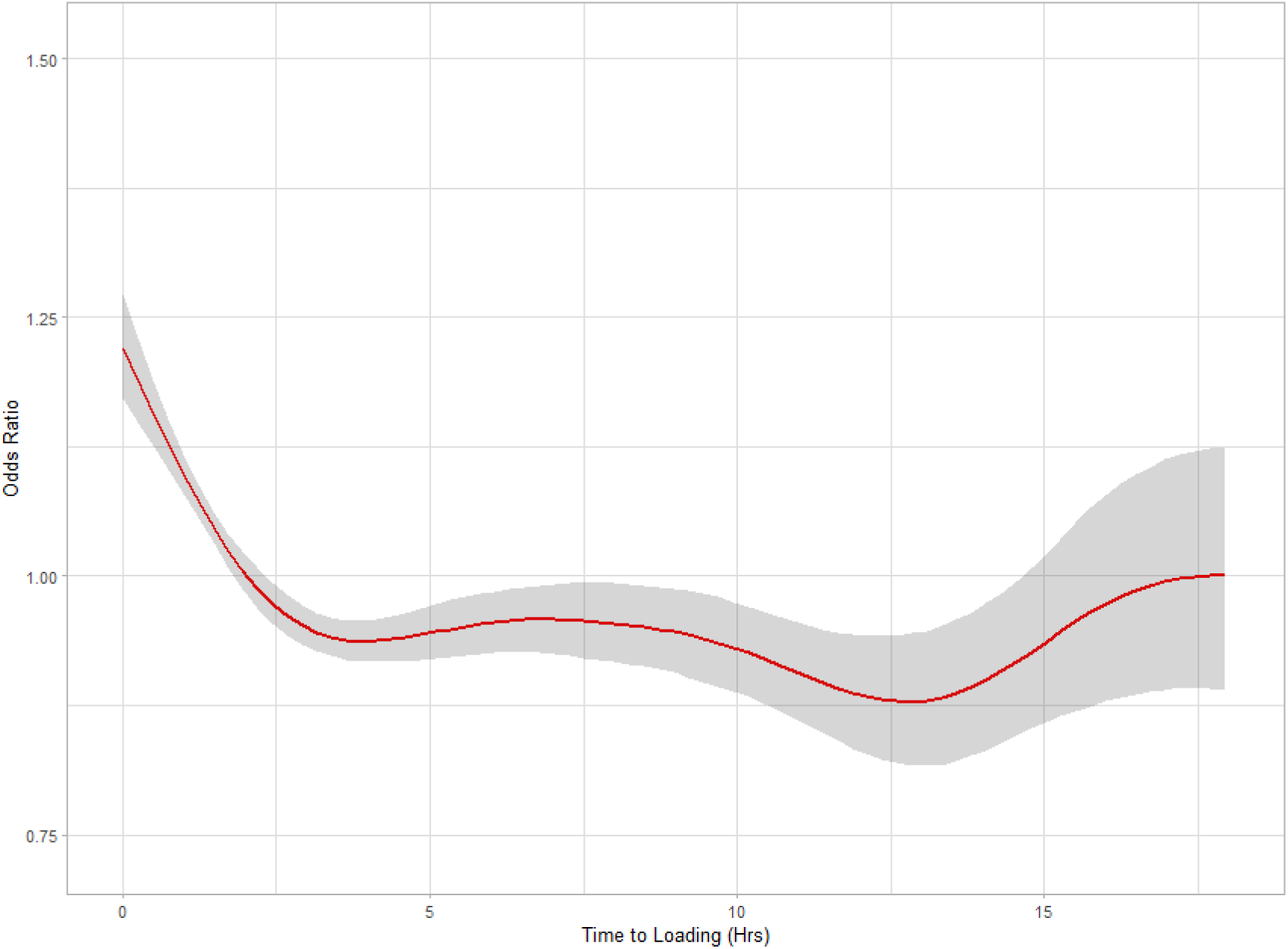
Generalized additive model plot of estimate against time to loading for growth with 95% credible intervals.

**Figure 3.**
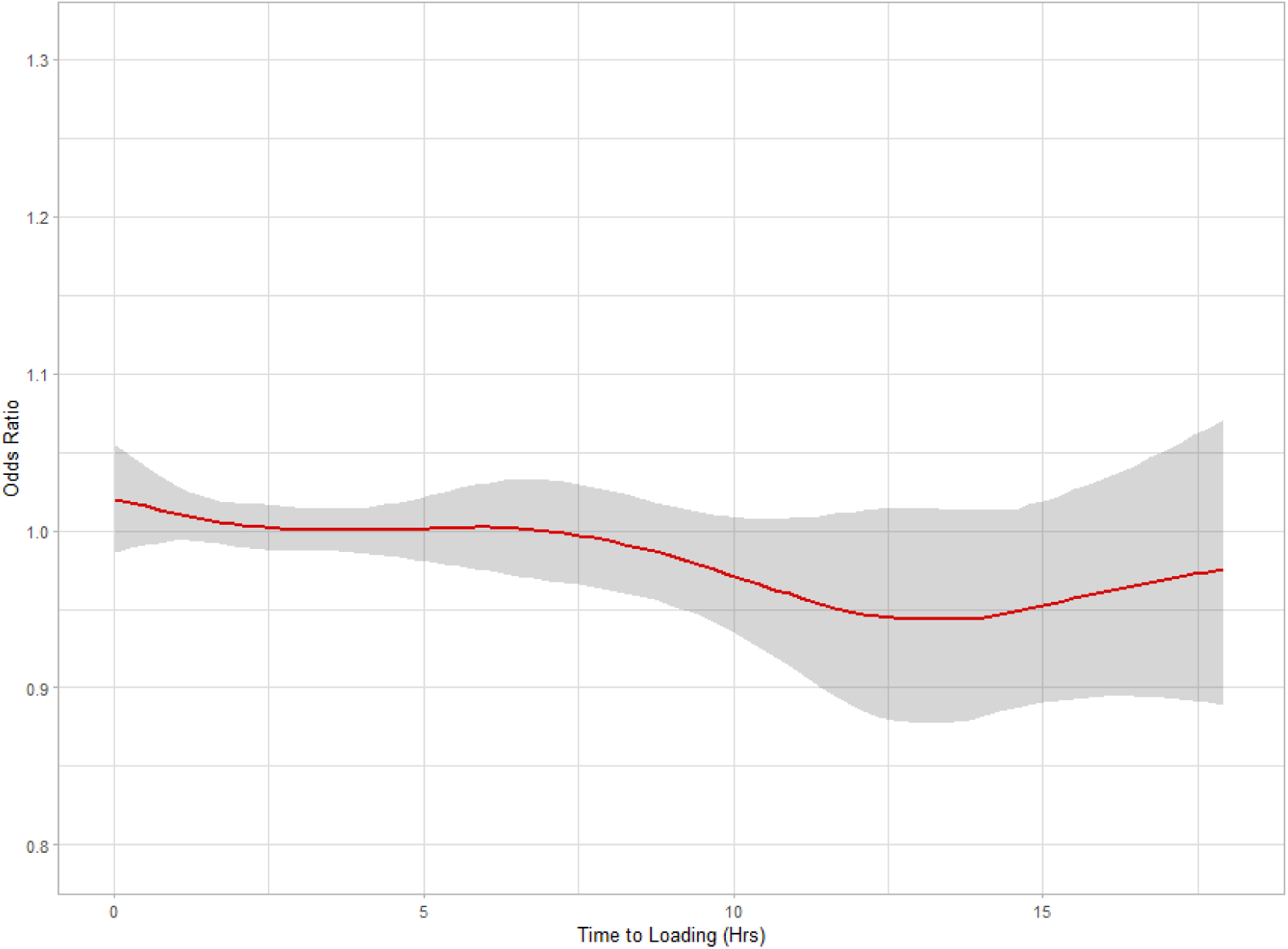
Generalized additive model plot of estimate against time to loading for growth with 95% credible intervals, adjusted for location, year, and working day.

**Figure 4.**
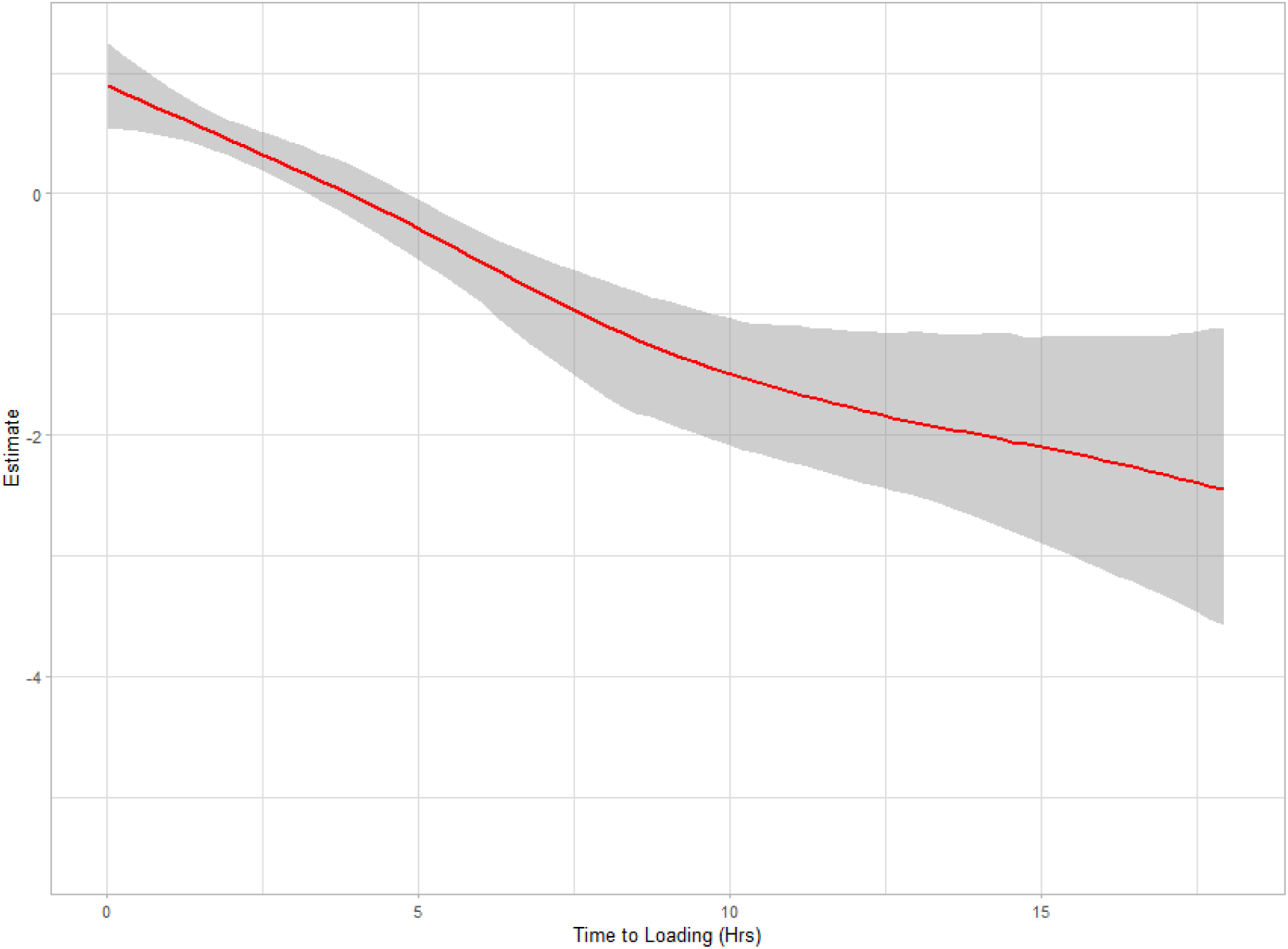
Generalized additive model of time to loading on time to detection with 95% credible intervals, without adjustment.

**Figure 5.**
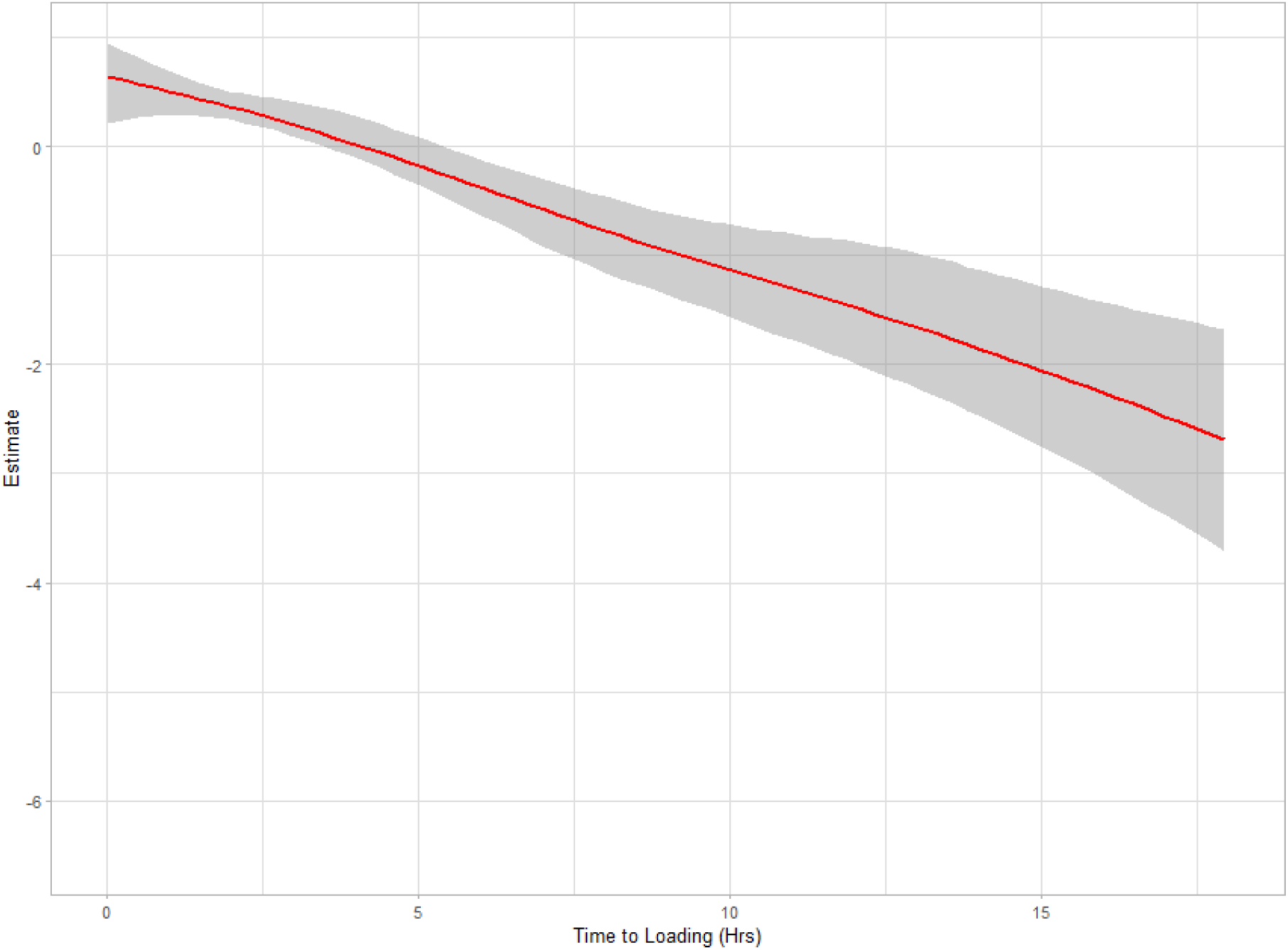
Generalized additive model of time to loading on time to detection with 95% credible intervals adjusted for location, working day, organism and year.

Considering specific groups of pathogens; the adjusted model for yeasts identified no effect (OR for growth from TTL was 1.01, 95% CrI 0.98 - 1.03), nor for anaerobes (OR 0.98, 95%CrI 0.95 – 1.00).

### Loading blood culture bottles quickly leads to proportionately less time on the incubator: reducing overall time to positivity in two ways

The unadjusted model identified that for every hour loaded earlier, there was an increase of 0.21 hours (95%CrI 0.16-0.27) of TTD. In the mixed effects model we saw a similar effect: 0.18 hours (95%CrI 0.13 – 0.24). This suggests that for every hour increase in time to loading there is 10.98 (95% CrI 7.8 – 13.2) minutes less TTD. As the total time to positivity is TTL + TTD, this means that some of the benefit of loading earlier is reduced by longer time to detection, and is consistent with bacterial growth occurring prior to loading, whilst in the culture set is in transit, but at a reduced rate. We did not identify any evidence of non-linearity, as expected.

### The effect on yield is dependent on the species of microorganisms: with a large effect on Streptococci including S. pyogenes

It is well established that different organisms have differing growth rates in culture, and it may be that the effect of delayed loading on yield is species specific. Table 2 shows the list of isolates with greater than 250 occurrences with the mean time to loading and time to detection, exclusive of contaminants. *Escherichia coli* was the most common pathogen.

**Table 2.** Table of the isolates occurring more than 250 times, exclusive of contaminants, with mean TTL, median TTL and number of isolates; ordered by ascending mean TTL.

| Organism | Mean TTL | Mean TTD | SD TTL | N |
| --- | --- | --- | --- | --- |
| ***Streptococcus pneumoniae*** | 2.85 | 11.75 | 2.52 | 760 |
| ***Streptococcus pyogenes*** | 2.88 | 11.24 | 2.72 | 352 |
| ***Streptococcus dysgalactiae*** | 2.97 | 10.78 | 2.96 | 457 |
| ***Streptococcus agalactiae*** | 3.31 | 10.49 | 2.83 | 469 |
| ***Escherichia coli*** | 3.34 | 13.93 | 2.84 | 6261 |
| ***Staphylococcus aureus*** | 3.34 | 19.93 | 2.70 | 3526 |
| ***Proteus mirabilis*** | 3.37 | 17.66 | 2.87 | 423 |
| ***Serratia marcescens*** | 3.38 | 17.72 | 2.69 | 267 |
| ***Pseudomonas aeruginosa*** | 3.44 | 21.07 | 2.82 | 678 |
| ***Enterococcus faecalis*** | 3.45 | 16.35 | 2.84 | 532 |
| ***Klebsiella oxytoca*** | 3.53 | 15.54 | 2.78 | 304 |
| ***Klebsiella pneumoniae*** | 3.55 | 15.29 | 2.85 | 1107 |
| ***Enterococcus faecium*** | 3.61 | 16.50 | 2.79 | 655 |
| ***Enterobacter cloacae complex*** | 3.79 | 14.62 | 2.79 | 440 |
| ***Streptococcus mitis/oralis*** | 3.87 | 15.77 | 2.97 | 283 |
| ***Enterobacterales*** | 4.11 | 14.21 | 2.81 | 317 |

As we know that at the organism is unknown at the time of the blood culture being taken, and that the laboratory management of blood cultures is generally independent of the clinical management, so we should expect the time to loading to be broadly similar for each species. However, Table 2 demonstrates subtle differences in mean TTL, *Streptococcus pneumoniae, dysgalactiea* and *pyogenes* have the quickest mean TTL.

As this difference is not due to earlier loading, this is likely due to decreased yield with longer TTL in some species. For example, if a species cannot survive for more than 8hrs in a culture set before being loaded, then we will only get positive results for this species when the TTL is < 8hrs, lowering the mean TTL. We therefore repeated our above analysis focusing on each pathogen using the same approach for these common pathogens.

Figure 8 shows a forest plot with 95% credible intervals for the most common isolates and their odds ratio of growth from the TTL. The organisms showed the strongest effect for TTL reducing yield were *Streptococcus pneumonia, agalactiea* and *pyogenes*.

**Figure 7.**
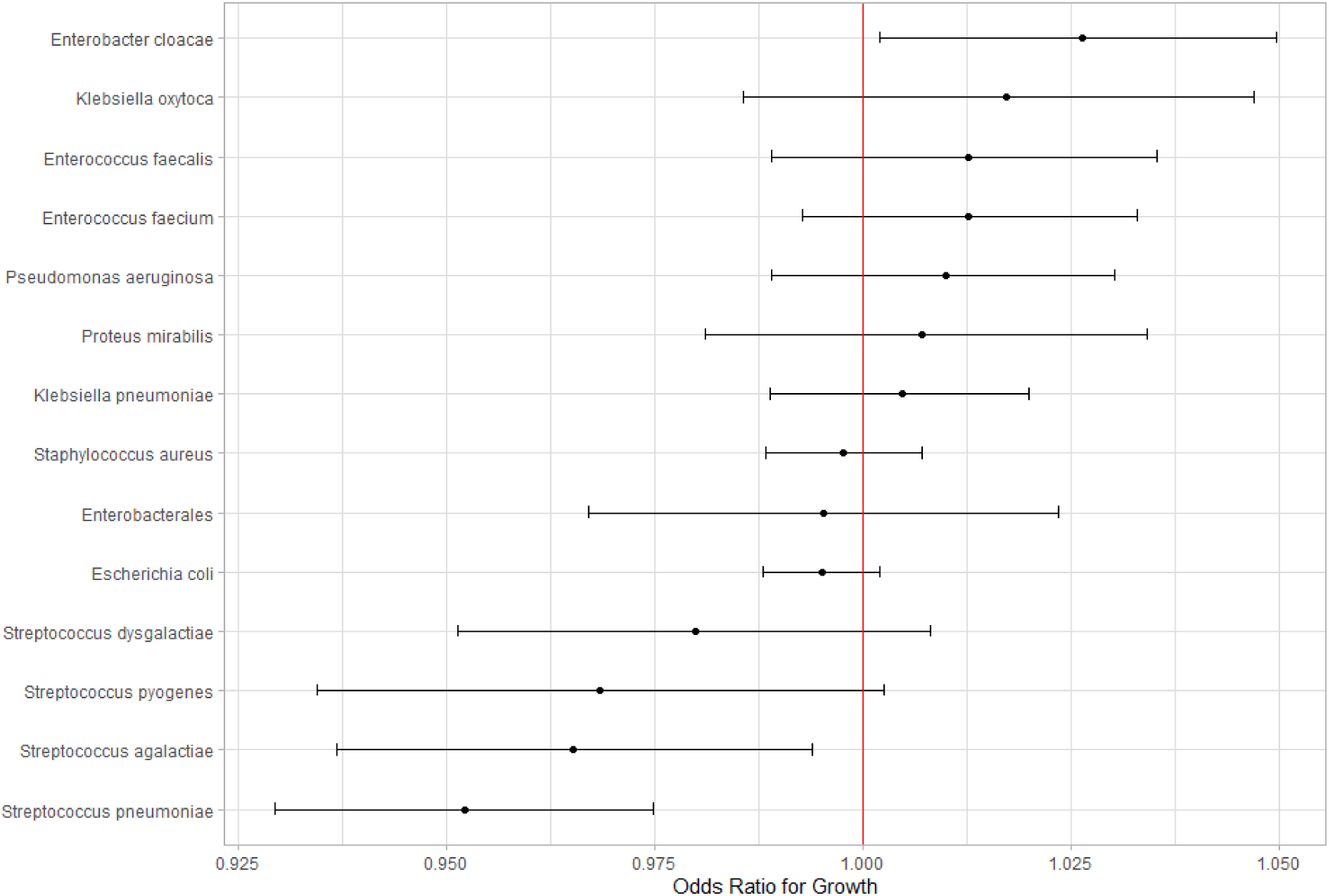
Forest plot for the estimate of growth per organism, adjusted for location, working day and year.

The beta haemolytic *streptococci* appear to be significantly affected by delays in loading, an odds ratio for *Streptococcus pyogenes* of 0.97 (95%CrI 0.93 – 1.0 per hour delay. This means that delaying loading *S. pyogenes* by 4 hours might lead to a loss of 11.4% of yield. *Streptococcus pneumonia* may be explained by the known phenomena of autolysis [15].

Figure 9 shows the relationship between the estimates on TTD and mean TTL for each organism listed in Table 2. Unsurprisingly, most isolates have figures below 0, meaning TTD is reduced for each hour they are delayed in getting to the laboratory, with the largest effects for the other *Enterobacterales* species and the beta haemolytic *Streptococci.* For all species, estimates were larger than 1: the bacterial growth before loading was never fast enough to mean that delayed loading had no effect on the overall time to positivity.

**Figure 9.**
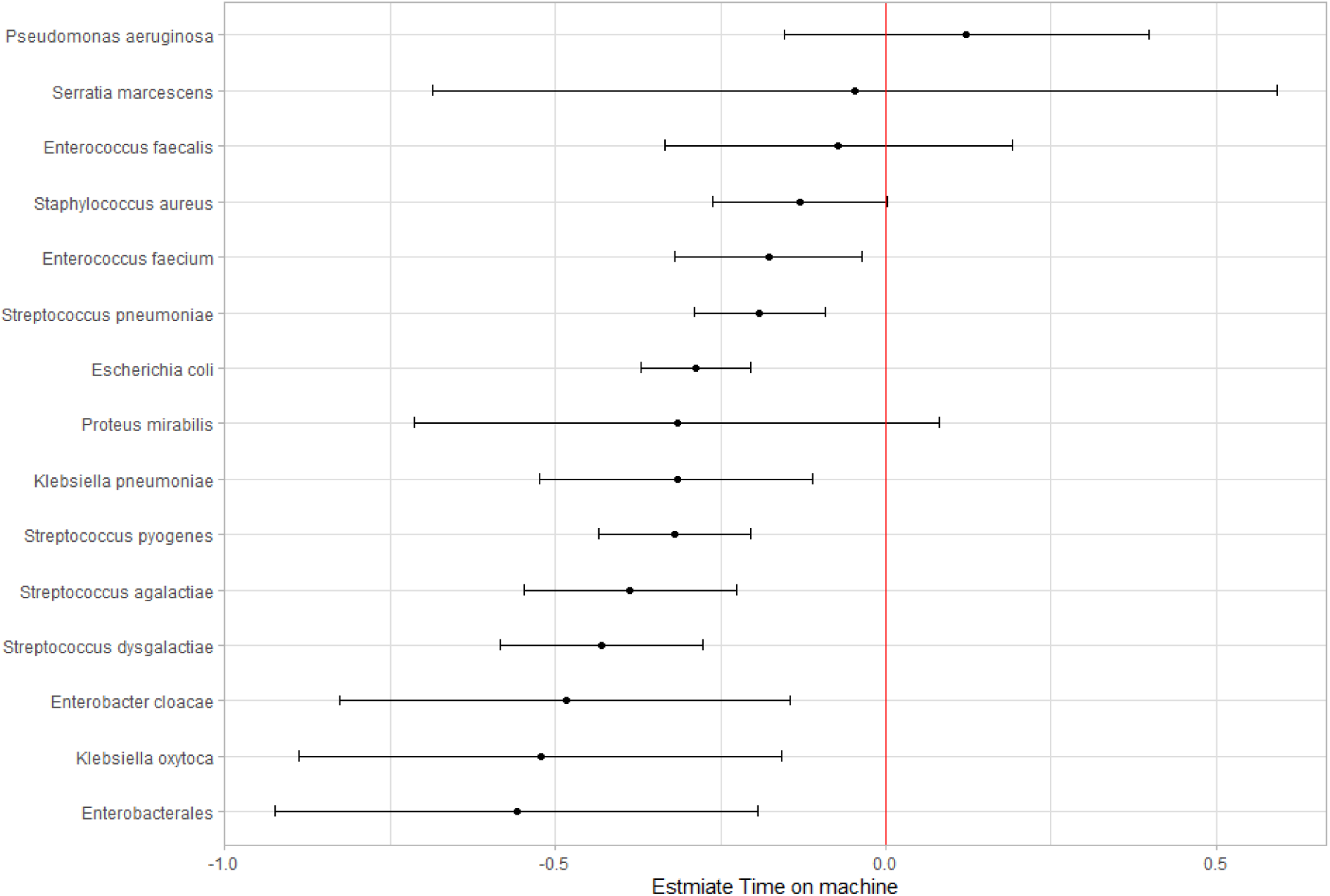
Forest plot of estimate on species time to detection from TTL.

## Discussion

This analysis of blood culture loading showed that a longer time to loading (TTL) was associated with a minimally lower rate of positivity, with an OR of 0.997 (95%CrI 0.994 - 1.001) for each hour. This was largely driven by loss of beta and alpha haemolytic *Streptococci*. The unique feature of *S pneumoniae* exhibiting autolysis may explain [15] its estimate and loss of growth within delayed blood culture bottles. The loss of *S pyogenes* is of both clinical and public health concern, a loss of up to 11.4% if cultures arrive in the laboratory at 4 hours from sampling. Invasive *S pyogenes* is a severe infection and can lead to seeding to distant sites, requiring prolonged antibiotics and multiple surgical interventions. Correctly diagnosing *Streptococcal* bacteraemia would reduce broad spectrum antimicrobial use, increase public health intervention and prevention, as well as improving diagnostic stewardship. In this the overall effect of earlier loading was small, suggesting that marginal benefit in overall yield would be expected by reducing the time to load. Whether the effect on delayed transport to the laboratory on yield is worth investing in infrastructure to reduce is debatable; it is well known that other factors have effects on yield that are far greater than that described here, principally blood volume [16].

### Comparisons with previous literature

Venturelli et al published an observational study on time to loading and positivity rates in 2017. They report the results of 50,955 culture results and found a similar effect (OR of 0.997, 95%CI:0.994-0.999) for growth for each hour in delay which matches our main result. However, they did not detect differences in sub-analysis of pathogens. Their laboratory only accepted culture sets in working hours but otherwise worked similarly our own laboratory [17]. A similar analysis from Canada noted that *Streptococcal species* were not recovered from blood cultures at the most distant sites, which had higher TTL. They found a similar OR for growth of 0.998 (95%CI 0.994 – 1.003) in logistic regression analysis [18].

Conversely, an interventional study of ICU patients’ blood culture sampling showed an increase in yield with increasing delay, 20.7% off site vs. 14.2% on site, p < 0.001 [19]. Their study had significantly fewer sets analysed at only 250 in each arm. Invitro studies have shown that time to loading at less than 24 hours has no decrease in yield, and that pre-loading temperature control at room temperature is also of no consequence [20]. Taken together these would suggest that there is, at best, a marginal improvement in yield with decreasing TTL.

The effect of TTL on time to positivity and TTD suggest that cultures spent less time on the incubators if they were loaded with a longer delay with a negative coefficient of –0.18, around 11 minutes quicker per hour delay. This is different to previous publications and the sources cited for the UK SMI where a coefficient of greater than 0 is described, suggesting that for every hour delay there is an increased TTD [21] [22]. This could be explained by the relatively short delay times here compared to the other studies, which looked at a difference of 12 hours or more between groups; or that the bottles here are subject to low level incubation whilst on the wards in our group that was not the case in the published groups, a factor that cannot be accounted for.

### Strengths and limitations

The strength of this model is that it accounts for different patient populations within the hospital by nesting wards within hospital sites in the multilevel Bayesian model, in this way we also account for the effect of baseline positivity areas being different distances to the laboratory, adjustment in this manner means that OR for growth dramatically falls to very little compared to the unadjusted analysis. One unaccounted for factor is that blood cultures are an imperfect test, and it is assumed in clinical practice that even negative blood cultures in an antibiotic naive patient may harbour pathogens. This means that we have no objective measurement for the presence of growth when looking at the various predictor variables, and as such the forest plots showing the estimate of growth from time to loading for the most common pathogens is relative to each other and not to an objective reference organism. This as an observational study and although we cannot rule out differences in loading time and practices that might affect yield, we cannot prove it.

## Conclusion

This analysis found that time to loading was independently and negatively correlated with a loss of yield at around 0.3% per hour, with a significant loss in some *Streptococci.* However, there is no clear evidence that this pre-analytic requirement affects yield for other pathogens.

## Funding

PW is funded by Medical Research Council grant MR/T005408/1. FH is funded by the NIHR Clinical Lectureship programme. For the purpose of open access, the author(s) has applied a Creative Commons Attribution (CC BY) licence to any Author Accepted Manuscript version arising from this submission.

## Data Availability

All data produced in the present study are available upon reasonable request to the authors

## Notes

### Competing Interest Statement

The authors have declared no competing interest.

