## Supplementary material 2 for "Less Haste, More Speed: Does delayed blood culture loading lead to adverse incubation time or yield?"

```
m1 <- brm(growth ~ time_to_load,
        data   = d_no_contams,
        warmup = 1000,
        iter   = 2000,
        chains = 4,
        cores = 4,
        threads = threading(4),
        family = bernoulli(link = "logit"),
        backend = "cmdstanr",
        stan_model_args = list(stanc_options = list("O1")))

m2 <- brm(growth ~ time_to_load + working_day + year + (1 | site/ward),
        data   = d_no_contams,
        warmup = 1000,
        iter   = 2000,
        chains = 4,
        cores = 4,
        threads = threading(4),
        family = bernoulli(link = "logit"),
        backend = "cmdstanr",
        stan_model_args = list(stanc_options = list("O1")))

m3 <- brm(growth ~ s(time_to_load),
        data   = d_no_contams,
        warmup = 1000,
        iter   = 2000,
        chains = 4,
        cores = 4,
        threads = threading(4),
        family = bernoulli(link = "logit"),
        backend = "cmdstanr",
        stan_model_args = list(stanc_options = list("O1")))

m4 <- brm(growth ~ s(time_to_load) + working_day + year + (1 | site/ward),
        data   = d_no_contams,
        warmup = 1000,
        iter   = 2000,
        chains = 4,
        cores = 4,
        threads = threading(4),
        family = bernoulli(link = "logit"),
        backend = "cmdstanr",
        stan_model_args = list(stanc_options = list("O1")))

m5 <- brm(time_on_machine ~ time_to_load,
        data   = d_only_pos,
        warmup = 1000,
        iter   = 2000,
        chains = 4,
        cores = 4,
```

```
          threads = threading(4),
          family = gaussian(),
          backend = "cmdstanr",
          stan_model_args = list(stanc_options = list("O1")))

m6 <- brm(time_on_machine ~ time_to_load + working_day + organism + (1 | site/ward),
          data   = d_only_pos,
          warmup = 1000,
          iter   = 2000,
          chains = 4,
          cores = 4,
          threads = threading(4),
          family = gaussian(),
          backend = "cmdstanr",
          stan_model_args = list(stanc_options = list("O1")))

m7 <- brm(time_on_machine ~ s(time_to_load),
          data   = d_only_pos,
          warmup = 1000,
          iter   = 2000,
          chains = 4,
          cores = 4,
          threads = threading(4),
          family = gaussian(),
          backend = "cmdstanr",
          stan_model_args = list(stanc_options = list("O1")))

m8 <- brm(time_on_machine ~ s(time_to_load) + working_day + organism + (1 | site/ward),
          data   = d_only_pos,
          warmup = 1000,
          iter   = 2000,
          chains = 4,
          cores = 4,
          threads = threading(4),
          family = gaussian(),
          backend = "cmdstanr",
          stan_model_args = list(stanc_options = list("O1")))

list <- d_no_contams |>
  count(organism, sort = T) |>
  filter(n > 300) |>
  pull(organism)

model_function <- function(organism_name) {

  d_working <- d_no_contams |>
    filter(organism == organism_name | is.na(organism))

  m_growth <- brm(growth ~ time_to_load + working_day + year + (1 | site/ward),
                  data   = d_working,
                  warmup = 1000,
                  iter   = 2000,
                  chains = 4,
                  cores = 4,
```

```
            threads = threading(4),
            family = bernoulli(link = "logit"),
            backend = "cmdstanr",
            stan_model_args = list(stanc_options = list("O1")))
}
map(list, model_function)
```
