## Supplementary material 1 for "Less Haste, More Speed: Does delayed blood culture loading lead to adverse incubation time or yield?"

List of organisms considered pathogenic:

Enterobacterales
Pseudomonadales
Enterococcus
Staphylococcus aureus
Streptococcus pneumoniae
Beta-hemolytic streptococcus
Streptococcus anginosus
Streptococcus gallolyticus
Haemophilus influenzae
Haemophilus parainfluenzae
Streptococcus constellatus
Stenotrophomonas maltophilia
Streptococcus intermedius
Neisseria meningitidis
Corynebacterium striatum
Aerococcus urinae
Pasteurella multocida
Finegoldia magna
